# Ethiopian Healthcare Workers’ Experiences During the COVID-19 Pandemic

**DOI:** 10.1101/2022.02.01.22270247

**Authors:** Averi Chakrabarti, Solomon Tessema Memirie, Delayehu Bekele, Mizan Kiros, Christina L. Meyer, Phuong N. Pham, Patrick Vinck, Stéphane Verguet

## Abstract

**Background:** The COVID-19 pandemic has caused widespread health and socioeconomic disruptions around the world. Understanding the impact that this crisis has had on health workers and the delivery of routine health care services within countries provides evidence on pandemic preparedness and response. Here, we conduct an investigation into these factors for the Ethiopian context.

**Methods and findings:** We conducted an online cross-sectional survey with Ethiopian health care professionals between August 27 and October 10, 2020 via existing research networks. The variables of interest were confidence in COVID-19 related knowledge, training and experience, the adoption of precautionary health practices, risk perceptions, and respondent concerns. The majority of surveyed health care professionals in Ethiopia reported seeing fewer patients than usual during the COVID-19 crisis, gaps in pandemic training, inadequate access to personal protective equipment (PPE) and barriers to accessing COVID-19 testing. Most health care professionals were also deeply concerned and worried about their own COVID-19 risks and the likelihood that they would transmit the disease to others.

**Conclusions:** Our study findings point to a possible reduction in routine health care services during the COVID-19 pandemic and gaps in pandemic preparedness in Ethiopia. The ministry of health and other stakeholders should work towards improving access to PPE and testing, and identify approaches to ensure that essential healthcare provision (such as immunizations) is not disrupted during crises akin to the COVID-19 outbreak.

## Introduction

Since the first report of the novel coronavirus disease 2019 (COVID-19) in December 2019, the number of confirmed cases and deaths have increased rapidly worldwide [1,2]. As of March 1, 2021, more than 117 million confirmed cases and 2.6 million deaths had been reported globally [2]. The pandemic has also led to widespread socioeconomic disruptions; estimates from the International Monetary Fund (IMF) suggest that COVID-19 will have caused 90 million people to experience extreme deprivation in 2020 [3].

Like other African countries, Ethiopia, the second most populated country on the continent, has experienced relatively low cases of COVID-19 [4]. Nevertheless, as of March 2021, the country experienced nearly 160,000 cases and over 2,360 deaths, making it among the most affected countries on the continent [2,4]. Furthermore, more than 2490 health care workers were admitted to a facility due to COVID-19, of whom 19 have died [5]. Negative repercussions due to the pandemic have harmed Ethiopia’s economic, social and educational sectors [3,6,30]. Future waves of COVID-19 in African countries, including Ethiopia (though less distinct due to limited and inconsistent testing volume in Ethiopia), could overwhelm the health system [7]. With just 77 physicians per 1,000,000 people, one of the lowest physician densities in the world, Ethiopia already faces critical challenges in healthcare services delivery [8].

The prevention and control measures employed to suppress and mitigate the health effects of the COVID-19 pandemic in Ethiopia and other low- and middle-income countries (LMICs) are expected to place immense pressure on health systems and compromise the delivery of essential health services to the public [9-11]. Worryingly, pandemic-driven decreases or delays in routine care seeking may come to surpass the pandemic’s direct health impact [12,13]. A modeling study assessing the pandemic’s potential impact on maternal, neonatal and child health services in LMICs estimated that there might be increments of 12,000-57,000 maternal deaths and 1.2 million child deaths [12]. Another study conducting a benefit-risk analysis of routine childhood immunization during the COVID-19 pandemic in Africa demonstrated that 3-84 deaths could be averted for every COVID-19 attributable death depending on the scenarios compared [13]. It is important to note here that as with previous disease outbreaks (such as Ebola in Africa), declines in health service receipt during COVID could be driven not only be supply side factors (e.g. staffing issues), but also by demand side problems (such as fear of contagion among patients) (31, 32).

Assessing the impact of the coronavirus pandemic on health systems could help breach the evidence gap on pandemic preparedness and response in LMICs, such as those in sub-Saharan Africa. In this paper, we surveyed how the work activities of physicians, nurses and other health care workers in Ethiopia have been shaped by the COVID-19 pandemic. Specifically, we seek to gauge how COVID-19 has impacted their work and how they have responded to changes in their work requirements during the pandemic. In the face of Ethiopia’s limited health workforce, understanding these personnel’s activities and circumstances of work could be vital to leverage human resources in the fight against the pandemic [8,14].

## Methods

We conducted a cross-sectional survey using a standardized self-administered questionnaire via KoBoToolbox, an online humanitarian data collection platform [15]. The questionnaire was developed by researchers with experience conducting surveys among healthcare workers and knowledge of Ethiopia’s healthcare system. The instrument included questions on changes in patient load, confidence in knowledge of COVID-19, trainings received, access to protective equipment, treatment of COVID-19 patients and personal concerns related to the pandemic: the detailed questionnaire is given in the supplementary webappendix. Anonymous response data from the completed surveys were stored on a secure server, accessible only by members of the study team. Data collection was conducted between August 29 and October 10, 2020.

### Study population

The targeted population comprised health care professionals in Ethiopia, including physicians, health officers, nurses, and midwives with access to email. Potential respondents were sent the URL link for the survey through health care professional associations, namely the email listservs for the Ethiopian Medical Association (EMA), the Ethiopian Pediatric Society (EPS), the Ethiopian Society of Obstetricians and Gynecologists (ESOG), the Ethiopian Midwifery Association (EMwA), the Emergency Surgery and Obstetrics (IESO) association and the Ethiopian Nurses Association (ENA). A follow-up email reminder to complete the survey was sent to all the association members during the course of the study.

### Analysis

We pursue descriptive statistical analyses of the data collected through the online survey questionnaire. Specifically, we depict means and proportions on a range of indicators such as patient load, confidence in COVID-19 specific knowledge and training and access to personal protective equipment (PPE) among other measures.

### Ethical consideration

Ethical clearance was acquired from both the Harvard T.H. Chan School of Public Health Institutional Review Board (IRB) (Protocol #IRB20-1109) and the IRB of Saint Paul’s Millennium Medical College, Addis Ababa. Individual consent was sought from potential respondents prior to online survey participation. All data and survey answers were collected anonymously.

## Results

A total of 62 individuals responded to the online survey. The average age of respondents was 34 years and most participants (85%) were male. Physicians accounted for almost half of the sample, and nurses and integrated emergency surgical officers constituted 34% of the sample. The survey reached medical professionals with a broad range of specializations such as obstetrics/gynecology, pediatrics, infectious diseases and cardiology. Half of the respondents worked in Addis Ababa and another 36% worked in the Southern Nations, Nationalities and People’s region (SNNP), and Oromia and Amhara regions. Finally, 78% of the health facilities that respondents worked in were either general, specialized or primary hospitals, and 85% of the facilities were public. We emphasize here that the current study is likely to be capturing the experiences of a selected section of Ethiopia’s medical professionals (e.g. those have good internet connectivity and those who work in the capital or other wealthy areas), not the overall Ethiopian experience.

The survey asked respondents how current workloads, as defined by the number of patients seen, compare to normal operations before the COVID-19 pandemic. As depicted in Figure 1, 63% of the sample indicated seeing fewer patients than usual. The main factors cited as likely reasons for this decline in caseload were patient fears of contracting COVID-19 within health facilities, reductions in the provision of non-essential services at health facilities, restrictions on mobility and lack of transportation options to and from health facilities/patient home. Those who reported an increase in their current workload listed follow-up care (not related to COVID-19) and COVID-19 related healthcare services as the main factors driving the observed uptick in demand. Regardless of how the pandemic had changed the total number of patients seen by the medical professionals in the study sample, these individuals continued to see patients: 57 out of the 62 respondents reported having provided direct medical care to hospital patients during the previous four weeks.

**Figure 1.**
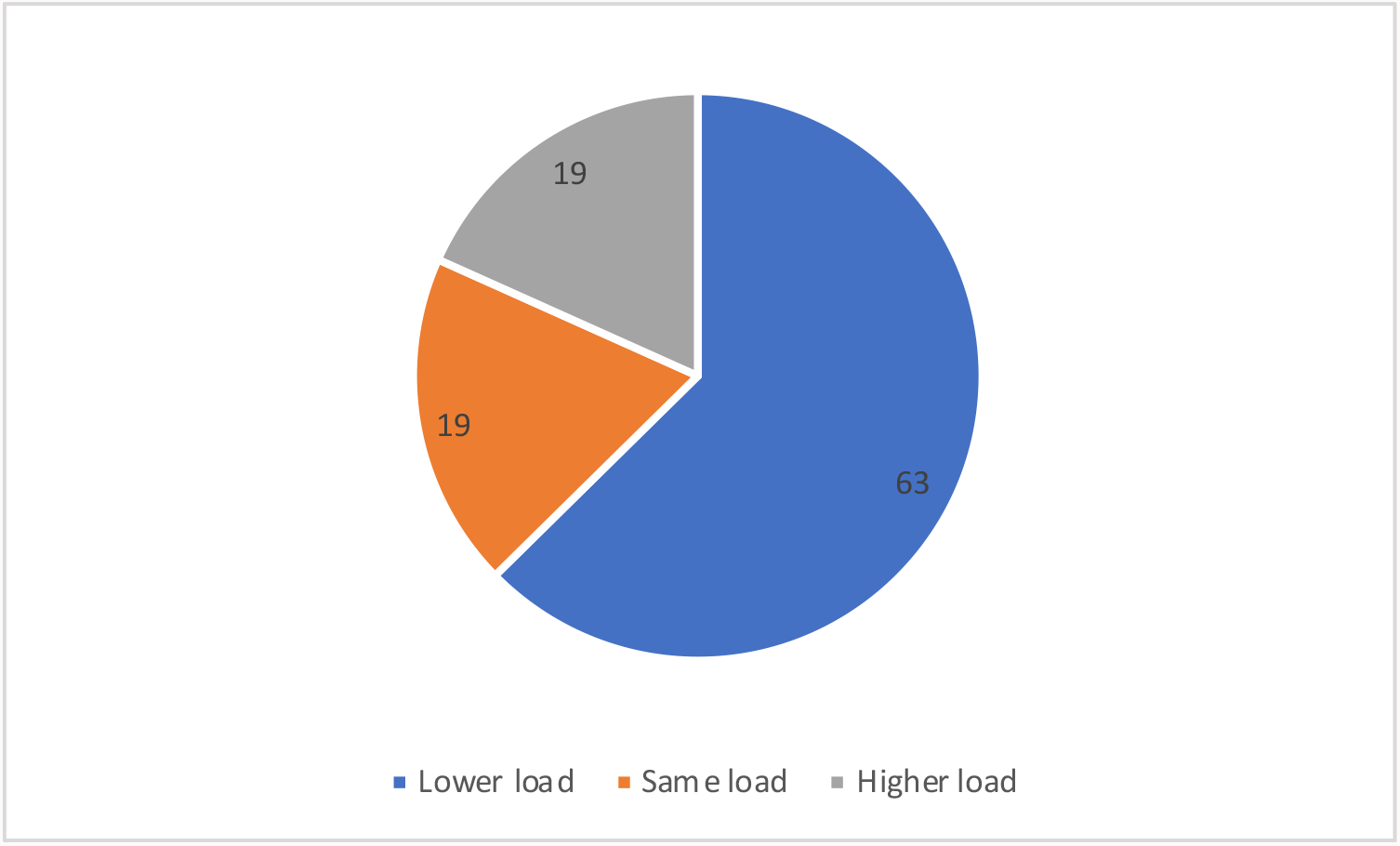
Distribution of how the current workload (number of patients seen) compared to normal operations before the COVID-19 pandemic (% reporting different categories)

Figure 2 conveys how much time respondents spent on COVID-19 activities such as training, seeing infected patients and administrative work. The reference period was the previous week. Close to one-fifth of the respondents were not carrying out any COVID-19 duties. A similar share of respondents spent most to all their time on COVID-19 related activities.

**Figure 2.**
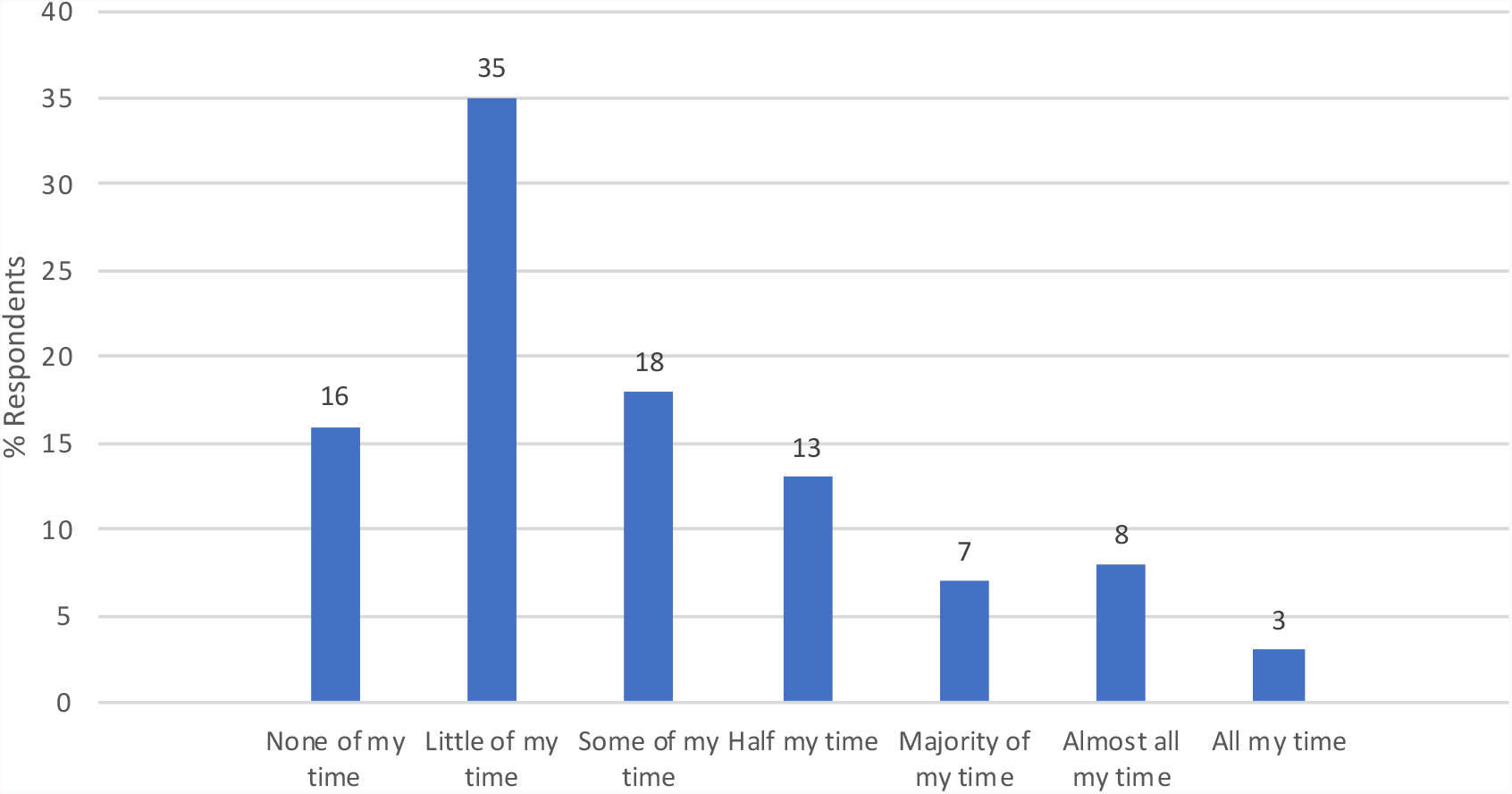
Time at work spent on COVID-19 activities during the previous week (% reporting different categories)

Several questions in the survey sought to gauge respondents’ confidence in knowledge of COVID-19 (Table 1). When asked to rate their overall level of information, 90% said they had an above-average level of understanding of the disease. Among individuals surveyed, 63% felt they were very able to explain COVID-19 to patients, community members or other individuals; 7% said they had low ability to do so. The survey asked about the areas in which medical professionals would prefer to have more information: the most commonly cited topics were protective measures for healthcare professionals, tests and diagnostics, and governmental response to/plan for COVID-19. The primary sources of information were scientific publications (35%), social media (23%), and television (15%).

**Table 1:** Confidence in knowledge of COVID-19 among surveyed Ethiopian health care professionals.

|  | Number<br>(%) |
| --- | --- |
| Overall level of information about COVID-19 |  |
| Very Good/Good | 56 (90) |
| Average | 4 (7) |
| Poor/Very Poor | 2 (3) |
| *Ability to explain COVID-19 to others* |  |
| Very able | 39 (63) |
| Somewhat able | 19 (31) |
| Little able | 4 (7) |
| *Primary source of information on COVID-19* |  |
| Scientific publications | 22 (35) |
| Social media | 14 (23) |
| Television | 9 (15) |
| Government communication | 8 (13) |
| Medical colleagues | 5 (8) |
| NGOs/UN agencies | 4 (7) |
| *Indicated the following were COVID-19 symptoms* |  |
| Cough, shortness of breath, difficulty breathing | 61 (98) |
| Fever, chills | 61 (98) |
| Loss of taste/smell | 54 (87) |
| Swelling of tongue, itchiness | 18 (29) |
| Nausea, vomiting, diarrhea | 49 (79) |
| Burning with urination | 7 (11) |
| Observations: n = 62. |  |

The survey assessed whether or not participants identified specific symptoms as being associated with COVID-19. Almost the entire sample was able to identify the most common symptoms— cough, shortness of breath, difficulty in breathing, fever and chills. A high proportion (87%) also pointed to the loss of taste/smell as symptoms. However, about one in three participants identified at least one incorrect symptom, such as burning while urinating or swelling of the tongue.

Table 2 contains statistics on respondents’ experiences with trainings related to COVID-19. Just under half the sample reported no exposure to such trainings/workshops. Of those who did attend any training, 58% reported that the last training received was more than two months before they responded to the survey, likely at the beginning of the pandemic, and 18% reported that the last training occurred during the previous month. Places of work provided the bulk of the trainings; government and non-governmental organizations are also frequent training organizers. Topics that appeared to be most commonly covered by COVID-19 trainings were disease transmission, symptoms, prevention and protective measures for healthcare professionals; issues that seemed to be least discussed were vaccinations, outreach and the government response. The information received at the workshops was “very useful” for 42% of attendees in improving care for patients and for 61% in protecting themselves from potential infection. The need for additional training remained high: 69% indicated that receiving training on COVID-19 would be very useful.

**Table 2:** COVID-19 training received among surveyed Ethiopian health care professionals.

|  | Number (%) |
| --- | --- |
| *Received/Participated in training or workshop on COVID-19* |  |
| None | 29 (47) |
| One training | 29 (47) |
| Multiple trainings | 4 (7) |
| *When was the last training? (of those receiving any training)* |  |
| More than two months ago | 36 (58) |
| Within the last two months ago | 15 (24) |
| Within the last month | 7 (12) |
| Within the last week | 4 (6) |
| *Who provided the training? (of those receiving any training)* |  |
| Place of practice/work | 28 (45) |
| Government | 17 (27) |
| NGO | 17 (27) |
| Other | 9 (15) |
| *How useful would COVID-19 training at this point be?* |  |
| Not at all useful | 2 (3) |
| Little useful | 4 (7) |
| Somewhat useful | 13 (21) |
| Very useful | 43 (69) |
| Observations: n = 62. |  |

We convey reports on the current availability of PPE, such as masks and gloves at places of work in Figure 3. Half the sample reported little availability of such PPE. It is worth pointing that almost half the sample indicated that PPE was little available even prior to COVID-19. Therefore, the reported low levels of availability do not represent a decline in the aftermath of the COVID-19 outbreak. Despite low availability from work, 86% of respondents said they were using PPE more frequently as compared to during normal operations (Figure 3).

**Figure 3.**
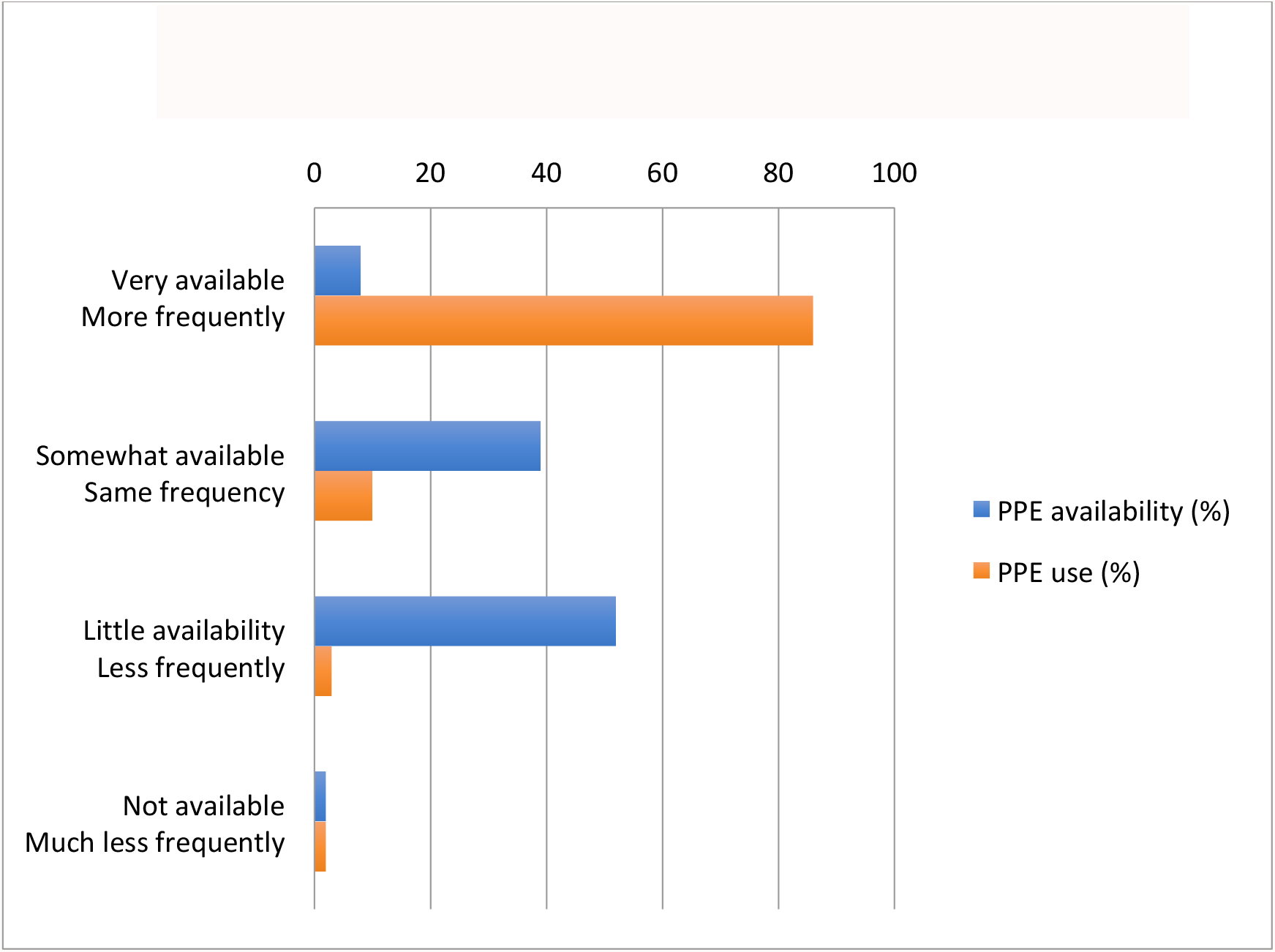
Current availability and use of protective equipment (% reporting different categories)

The survey posed several questions specifically related to face masks (Table 3). Of all respondents, 73% reported wearing masks at all times when seeing patients and another 19% reported wearing masks most of the time. Of those who said that they did not wear masks consistently, the majority indicated this was because of low availability. Respondents were asked to report where they had obtained masks for work: the most common sources appeared to be places of employment and personal purchases. Only 5% reported getting masks from their workplace on a daily basis; 31% received these weekly and 26% had only irregular access to masks at work.

**Table 3:** Protective equipment – face masks – used by surveyed Ethiopian healthcare professionals.

|  | Number (%) |
| --- | --- |
| *Frequency of wearing masks when seeing patients currently* |  |
| All the time | 45 (73) |
| Most of the time | 12 (19) |
| Some of the time | 3 (5) |
| Never see patients | 2 (3) |
| *Frequency with which new face masks are obtained from place of work* |  |
| Never | 4 (7) |
| Irregularly | 16 (26) |
| Monthly | 10 (16) |
| Weekly | 19 (31) |
| Every other day | 10 (16) |
| Every day | 3 (5) |

Observations: n = 62.

In Table 4, we present statistics on the respondents’ interactions with potential COVID-19-positive patients. In the sample, 63% indicated that they currently saw or interacted with patients potentially infected with COVID-19. Of those who responded in the affirmative, just 25% reported that patients wore masks all the time. Overall, 27% of respondents said they felt very able to detect potential cases of COVID-19; about three-quarters of the surveyed professionals had seen patients with symptoms consistent with COVID-19 and 68% had sought COVID-19 testing for patients. The most common barriers for accessing tests were the unavailability of testing supplies, certain patients not being prioritized by current testing policies and waiting times.

**Table 4:** Interactions of surveyed Ethiopian healthcare professionals with patients.

|  | Number (%) |
| --- | --- |
| *Currently sees/interacts with patients potentially infected with coronavirus* |  |
| Yes | 39 (63) |
| No | 23 (37) |
| *Frequency with which patients wear masks*<br>*(of those who currently see/interact with potentially infected patients)* |  |
| All the time | 16 (25) |
| Most of the time | 24 (39) |
| Some of the time | 20 (33) |
| Rarely | 2 (3) |
| *Perceived ability to detect potential COVID-19 cases among patients* |  |
| Very able | 17 (27) |
| Somewhat able | 36 (58) |
| Little able | 8 (13) |
| Not at all able | 1 (2) |
| *Has seen/interacted with patients showing COVID-19 symptoms* |  |
| Yes | 46 (74) |
| No | 16 (26) |
| *Has personally sought to obtain testing for patients showing COVID-19 symptoms* |  |
| Yes | 42 (68) |
| No | 20 (32) |

Observations: n = 62.

In Table 5, we present data on respondents’ concerns for themselves. Almost three-quarters of the respondents felt much at risk of contracting the disease. Concerns about transmitting the disease to patients and hospital visitors (friends and family) occurred among 96% of those surveyed. However, about 88% of respondents felt that they were very or somewhat able to help reduce the spread of COVID-19 in their communities. The surveyed medical professionals experienced high levels of stress, with 50% feeling nervous or anxious several days during the past two weeks and 44% experiencing constant worrying with the same frequency. A fourth of the sample reported facing abuse on account of being a medical professional during the pandemic. The most common abuse involved insults or being forced out of housing.

**Table 5:** Concerns among surveyed Ethiopian healthcare professionals.

|  | Number (%) |
| --- | --- |
| *Perceptions of own level of risk* |  |
| Very much at risk | 42 (68) |
| Somewhat at risk | 18 (29) |
| Little risk | 2 (3) |
| *Concerned about transmitting COVID-19 to patients/hospital visitors* |  |
| Very concerned | 36 (58) |
| Somewhat concerned | 21 (34) |
| Little concerned | 5 (8) |
| *Concerned about transmitting COVID-19 to friends/family* |  |
| Very concerned | 40 (65) |
| Somewhat concerned | 19 (31) |
| Little concerned | 3 (5) |
| *Perception of contribution towards reducing the spread of COVID-19 in the community* |  |
| Very able | 22 (35) |
| Somewhat able | 33 (53) |
| Little able | 7 (12) |
| *Frequency of feeling nervous, anxious or on edge over the last two weeks* |  |
| Nearly every day | 7 (11) |
| More than half of the days | 10 (16) |
| Several days | 31 (50) |
| Not at all | 14 (23) |
| *Frequency of being bothered by not being able to stop or control worrying over the last two weeks* |  |
| Nearly every day | 6 (10) |
| More than half of the days | 9 (15) |
| Several days | 27 (44) |
| Not at all | 20 (32) |
| *Has experienced anger/verbal or other abuse because of profession during the pandemic* |  |
| Yes | 15 (24) |
| No | 47 (76) |

Observations: n = 62.

## Discussion

This paper reports results from an online survey administered to health care professionals in Ethiopia between August and October 2020 to gauge their experiences during the COVID-19 pandemic. A sample of 62 individuals responded to questions on changes in patient load, confidence in knowledge of COVID-19, trainings received, access to protective equipment, treatment of COVID-19 patients, and personal concerns related to the pandemic.

Overall, most health care professionals reported seeing fewer patients than usual, a trend largely attributed to patient fears of contracting COVID-19 at health facilities (which led them not to visit health facilities), reductions in non-essential health service provision, and restrictions in mobility in the country. This finding is consistent with reports from various other countries that point to declines in outpatient care and hospitalization during the COVID-19 pandemic, both for emergency conditions as well as elective procedures [9-11,16-18]. Frequently cited barriers in these other settings also include patient fears, cancelation/reduction of certain services, and difficulties in reaching health facilities [16-18]. Comparison of routine health services utilization data, pre and during the pandemic in Ethiopia, showed a sharp decline in service utilizations during the early period of the pandemic especially for preventive services [19]. To address the potential fallout of reductions in careseeking, policymakers should employ a clear communication strategy to highlight what services are available at health facilities, and the infection prevention and control measures adopted at these centers. Approaches besides face-to-face contact, such as remote consulting via mobile phones, should also be considered.

In order to effectively respond to the COVID-19 pandemic, adequate knowledge and preparedness regarding COVID-19 transmission, and prevention and management by health workers are critical [20-23]. Even though the majority of the current study’s sample (90%) felt comfortable about their level of information (a comparable finding was reported in an other study in Ethiopia) [24], almost half had not received any training on COVID-19 and a large proportion (69%) considered additional COVID-19 training at this point to be very useful. These trends are mirrored in findings from Libya that demonstrated low levels of training on COVID-19 among health care workers and therefore an overwhelming reliance on social media as major sources of information [21]. Such training gaps can have significant negative implications since knowledge is crucial for promoting the necessary behavioural changes for prevention and control such as early diagnosis and isolation of cases [25].

The starkest indication of Ethiopia’s inadequate pandemic preparedness was that more than half the respondents indicated that PPE (e.g. masks and gloves) were inadequately available, and only 5% obtained face masks on a daily basis from their workplace. Despite such shortages, medical professionals appeared to secure access to some PPE: 86% of respondents said they are wearing PPE more frequently than before the pandemic.

About two-thirds of respondents saw patients potentially infected with COVID-19 and had personally sought testing for their patients. The most commonly reported barriers in accessing testing were the unavailability of testing supplies, patients not meeting testing priorities and waiting times. It is worth pointing that COVID-19 reverse transcription-Polymerase chain reaction (RT-PCR) testing capacity has improved substantially over time in Ethiopia [26]. As of the beginning of December, 2020, nearly 1.7 million individuals had already been tested, with total per capita testing of nearly 15,000 per 1 million population [27]. Recent statistics, however, showed a decline in the number of tests and an increase in the positivity rate [27].

Our results also highlight that Ethiopia’s medical professionals are deeply concerned about their own COVID-19 risks and the likelihood that they could transmit the disease to others. Since the risk of acquiring infection and reporting a positive test is higher among frontline health care workers [28], ensuring the safety of medical personnel is essential to providing the best care possible for infected individuals [23]. Additionally, non-trivial proportions of the study sample reported feeling anxious and worried, factors that are likely to have negative consequences on health personnel’s mental health [29]. Further studies on the mental health impact of COVID-19 and designing a coping strategy accordingly is recommended.

The generalizability of our study findings is limited by the sample size—62 health care professionals completed the survey. Most respondents (88%) were from Addis Ababa, SNNP, Oromia and Amhara regions and we received few/no responses from regions such as Somali, Gambella, Afar and Benishangul-Gumuz. Low access to internet and the relatively limited experience of potential respondents to online surveys in those areas are possible reasons for these low response rate. More than half of the respondents were from the capital city, Addis Ababa, where internet access is better than in other parts of the country. The capital city is also the place that is most impacted by COVID-19 and where most of the COVID-19 cases are found. Another limitation of the study is the use of the English language for the survey rather than the local dialects.

Despite these shortcomings, our study provides a glimpse of Ethiopia’s health system preparedness and the healthcare workers’ experiences during the COVID-19 pandemic. Specifically, the findings highlight specific steps that Ethiopia would need to take to best leverage its health care workers’ abilities to most effectively tackle the COVID-19 (or a future) pandemic in the country. These steps include providing targeted training, ensuring access to PPE for frontline workers, improving access to COVID-19 testing, and equipping health professionals with the coping strategies they need to deal with potential mental health ramifications.

## Supporting information

Supplemental survey tool

## Data Availability

All data produced in the present study are available upon reasonable request to the authors.

## Acknowledgements

We are grateful to all survey participants and their respective professional societies.We thank the Maternal and Child Health Directorate, Federal Ministry of Health, Ethiopia and its participants to a vaccine economics webinar (January 2021) for valuable comments.

## Funding

We acknowledge funding support from Gavi the Vaccine Alliance, the Bill & Melinda Gates Foundation (INV-101074), and Bergen Center for Ethics and Priority Setting (BCEPS). The funder of the study had no role in study design, data collection and analysis, decision to publish, or preparation of the manuscript. The corresponding author had full access to all the data in the study and had final responsibility for the decision to submit for publication.

## Author Contributions

STM and SV initiated the study. AC, CLM, DB, PNP, PV, STM, and SV prepared the data collection tool. PV prepared the online platform. AC did the analysis. STM wrote the first draft of the manuscript. All authors contributed to the writing and editing of the manuscript.

## Competing interests

The authors declare that they have no competing interests.

## Abbreviations

COVID-19: coronavirus disease-2019
LMIC: low- and middle-income countries
PPE: personal protective equipment
SNNP: Southern Nations, Nationalities and People’s.

## Notes

### Competing Interest Statement

The authors have declared no competing interest.

### Author Declarations

IRB of Harvard T.H. Chan School of Public Health (Protocol #IRB20-1109) and IRB of Saint Paul Millennium Medical College gave ethical approval for this work.

## References

1. Huang C, Wang Y, Li X, et al. Clinical features of patients infected with 2019 novel coronavirus in Wuhan, China. Lancet 2020;395:497–506. doi:10.1016/S0140-6736(20)30183-5.

2. Johns Hopkins University. Coronavirus resource center. 29/10/2020. https://coronavirus.jhu.edu/.

3. International Monetary Fund. 2020. World Economic Outlook: A Long and Difficult Ascent. Washington, DC, October.

4. Mbow M, Lell B, Jochems SP, et al. COVID-19 in Africa: Dampening the storm? The dampened course of COVID-19 in Africa might reveal innovative solutions. Science 2020; 3696504:624–627.

5. Federal Ministry of Health Ethiopia. Daily report: case management and facility readiness section-Personal communication. Addis Ababa, Ethiopia: Federal Ministry of Health; 2021.

6. WHO-Ethiopia. First case of COVID-19 confirmed in Ethiopia. 29/10/2020. https://www.afro.who.int/news/first-case-covid-19-confirmed-ethiopia.

7. Africa CDC. 2021. Outbreak brief #51: Coronavirus Disease 2019 (COVID-19) pandemic. 13/01/2021. https://africacdc.org/download/outbreak-brief-51-coronavirus-disease-2019-covid-19-pandemic/

8. The World Bank. Physicians (per 1,000 people) – Ethiopia. 18/01/2021. https://data.worldbank.org/indicator/SH.MED.PHYS.ZS?locations=ET.

9. Livingston EH. Surgery in a time of uncertainty: a need for universal respiratory precautions in the operating room. JAMA 2020;E1–E2.

10. Kamrath C, Monkemoller K, Biester T, et al. Ketoacidosis in children and adolescents with newly diagnosed type 1 diabetes during the COVID-19 pandemic in Germany. JAMA 2020; E1–E3.

11. Kansagra AP, Goyal MS, Hamilton S, Albers GW. Collateral effect of Covid-19 on stroke evaluation in the United States. The New England Journal of Medicine 2020. DOI: 10.1056/NEJMc2014816.

12. Roberton T, Carter ED, Chou VB, et al. Early estimates of the indirect effects of the COVID-19 pandemic on maternal and child mortality in low-income and middle-income countries: a modeling study. Lancet Glob Health 2020. https://doi.org/10.1016/S2214-109X(20)30229-1.

13. Abbas K, Procter S, Zandvoort KV, et al. Routine childhood immunization during the COVID-19 pandemic in Africa: a benefit-risk analysis of health benefits versus excess risk of SARS-CoV-2-infection. Lancet Glob Health 2020. https://doi.org/10.1016/S2214-109X(20)30308-9.

14. Mann C, Dessie E, Adugna M, and Berman P. 2016. Measuring efficiency of public primary hospitals in Ethiopia. Harvard T.H. Chan School of Public Health and Federal Democratic Republic of Ethiopia Ministry of Health. Boston, Massachusetts and Addis Ababa, Ethiopia.

15. KoBoToolbox -Data Collection Tools for Challenging Environments [Internet]. KoBoToolbox. 2021 Available from: https://www.kobotoolbox.org/

16. Rosenbaum L. The untold toll—the pandemic’s effects on patients without Covid-19. N Engl J Med. Published online April 17, 2020. doi:10.1056/NEJMms2009984.

17. Baum A, Schwartz MD. Admissions to Veterans Affairs hospitals for emergency conditions during the COVID-19 pandemic. JAMA. 2020;324(1):96–99. doi:10.1001/jama.2020.9972.

18. Ahmed SAKS, Ajisol M, Azeem K, et al. Impact of the societal response to COVID-19 on access to healthcare for non-COVID-19 health issues in slum communities of Bangladesh, Kenya, Nigeria and Pakistan: results of pre-COVID and COVID-19 lockdown stakeholder engagements. BMJ Global Health. 2020;5:e003042. doi:10.1136/bmjgh-2020-003042.

19. Harvard T.H. Chan School of Public Health. Ethiopia Dashboard. Available at: https://hsph.me/hs-performance-covid-eth. 26/02/2021.

20. WHO, 2020. Critical Preparedness, Readiness and Response Actions for COVID-19. Geneva, Switzerland: World Health Organization. Available at: https://apps.who.int/iris/bitstream/handle/10665/331511/Critical%20preparedness%20readiness%20and%20response%20actions%20COVID-10%202020-03-22_FINAL-eng.pdf?sequence=1&isAllowed=y. 14/12/2020.

21. Elhadi M, Msherghi A, Alkeelani M, et al. Assessment of healthcare workers’ levels of preparedeness and awareness regarding COVID-19 infection in low-resource settings. Am J. Trop. Med. Hyg. 2020;103(2):828–833.

22. Zhang M, Zhou M, Tang F, et al. Knowledge, attitude and practice regarding COVID-19 among healthcare workers in Henan, China. J Hosp Infect. 2020; 105: 183–187.

23. Huh S. How to train the health personnel for protecting themselves from SARS-CoV-2 (novel coronavirus) infection when caring for a patient or suspected case. J Educ Eval Health Prof. 2020; 17: 10. doi: 10.3352/jeehp.2020.17.10.

24. Asemahegn MA. Factors determining the knowledge and prevention practice of healthcare workers towards COVID-19 in Amhara region, Ethiopia: a cross-sectional survey. Tropical Medicine and Health. 2020;48:72. https://doi.org/10.1186/s41182-020-00254-3.

25. McEachan R, Taylor N, Harrison R, et al. Meta-analysis of the reasoned action approach (RAA) to understanding health behaviors. Ann Behav Med 2016;50: 592e612.

26. Mohammed H, Oljira L, Roba KT, et al. Containment of COVID-19 in Ethiopia and implication for tuberculosis care and research. BMC Infectious Diseases of Poverty. 2020; 9(131). https://doi.org/10.1186/s40249-020-00753-9.

27. Ministry of Health Ethiopia and Ethiopian Public Health Institute. COVID-19 reported cases in Ethiopia. https://twitter.com/lia_tadesse/status/1338888497864839168/photo/2.

28. Nguyen LH, Drew DA, Graham MS, et al. Risk of COVID-19 among front-line health-care workers and the general community: a prospective cohort study. Lancet Public Health. 2020;S:e475–83.

29. Stijelja S, Mishara BL. COVID-19 and psychological distress-changes in internet searches for mental health issues in New York during the pandemic. JAMA Internal Medicine. 2020; E1–E3.

30. Wieser, C., Ambel, A. A., Bundervoet, T., & Haile, A. (2020). Monitoring COVID-19 Impacts on Households in Ethiopia. https://openknowledge.worldbank.org/bitstream/handle/10986/33824/Results-from-a-High-Frequency-Phone-Survey-of-Households.pdf?sequence=5

31. Bali, S., Stewart, K. A., & Pate, M. A. (2016). Long shadow of fear in an epidemic: fearonomic effects of Ebola on the private sector in Nigeria. BMJ global health, 1(3), e000111.

32. Morse, B., Grépin, K. A., Blair, R. A., & Tsai, L. (2016). Patterns of demand for non-Ebola health services during and after the Ebola outbreak: panel survey evidence from Monrovia, Liberia. BMJ global health, 1(1), e000007.

