## Supplemental survey tool for "Ethiopian Healthcare Workers’ Experiences During the COVID-19 Pandemic"

### GENERAL INFORMATION

#### » GENERAL INFORMATION – RESPONDENT

GR01 - WHAT IS YOUR AGE? *

GR02 - WHAT IS YOUR GENDER? *

- ◯ Female
- ◯ Male
- ◯ Prefer not to say

GR03 - WHAT IS YOUR PRIMARY POSITION / ROLE? *

- ◯ Physician (all specialty)
- ◯ Resident
- ◯ Nurse
- ◯ Midwife
- ◯ Laboratory
- ◯ Health officer
- ◯ Administration
- ◯ Hospital leadership
- ◯ Other, specify

OTHER ROLE, SPECIFY

GR04 - WHAT IS YOUR SPECIALTY, IF ANY? *

- ◯ None / no clinical role
- ◯ General practitioner
- ◯ Anesthesiology
- ◯ Cardiology
- ◯ Dermatology
- ◯ Emergency Medicine
- ◯ Infectious Diseases
- ◯ Internal Medicine
- ◯ Obstetrician/Gynecologist
- ◯ Orthopedics
- ◯ Ophthalmology
- ◯ Pediatrics

#### ACTIVITY

A01 - HOW DOES YOUR CURRENT WORKLOAD (NUMBER OF PATIENTS SEEN) COMPARE TO NORMAL OPERATIONS BEFORE THE COVID-19 PANDEMIC? *

- Extremely lower load (<25%)
- Much lower load (<50%)
- Lower load (<75%)
- Somewhat lower load (<100%)
- Same load (100%)
- Somewhat higher load (>100%)
- Higher load (>125%)
- Much higer load (>150%)
- Extremely higher load (>175%)

A02 - WHAT WERE THE ISSUES THAT MOST CREATED THIS INCREASE IN VOLUME, IN YOUR OPINION? *

*multiple responses accepted*

- not related to health issues
- Regular cold
- Diarrhea
- COVID-19 symptoms
- Seeking test for COVID-19
- Preventive care (non-COVID)
- Follow-up care (non-COVID)
- Body aches
- Headache
- Free healthcare services
- Other, specify

OTHER ISSUES, SPECIFY

A03 - WHY DO YOU THINK YOU HAD FEWER PATIENTS COMPARED TO BEFORE THE PANDEMIC? *

*multiple responses accepted*

- People don't consult because they are afraid of contracting COVID-19 at facility
- Lack of transportation
- Other restriction on mobility
- Hours of operations at health facility are limited

#### INFORMATION

IN01 - HOW DO YOU JUDGE YOUR OVERALL LEVEL OF INFORMATION ABOUT COVID-19? *

Very poor — Poor — Average — Good — Very good

IN02 - AND HOW DO YOU JUDGE YOUR ABILITY TO EXPLAIN COVID-19 TO OTHERS LIKE PATIENTS OR MEMBERS OF THE COMMUNITY? *

Not at all able — Little able — Somewhat able — Very able

IN03 - IN WHICH OF THE FOLLOWING AREAS WOULD YOU LIKE TO HAVE MORE INFORMATION? *

*multiple responses accepted*

- [ ] Transmission
- [ ] Symptoms
- [ ] Tests / diagnostic
- [ ] Isolation / quarantine
- [ ] Prevention
- [ ] Care for patients
- [ ] COVID-19 treatments centers
- [ ] Outreach
- [ ] Protective measures for healthcare professionals
- [ ] Government response / plan
- [ ] Other, specify

OTHER TOPICS, SPECIFY

##### » Source ranking

**IN04 - BY ORDER OF IMPORTANCE, WHAT ARE YOUR MAIN SOURCES OF INFORMATION ABOUT COVID-19? PLEASE RANK YOUR TOP THREE MAIN SOURCES.

1ST SOURCE *

- Medical colleagues
- Government communication
- NGOs / UN agencies
- Relatives and friends
- Newspaper
- Television
- Radio
- Social media
- Other media
- Scientific publications
- Other source, specify

#### TRAINING

T01 - HAVE YOU PARTICIPATED IN OR RECEIVED A TRAINING / WORKSHOP FOR HEALTHCARE PROFESSIONALS RELATED TO COVID-19? *

- No
- Yes, one training
- Yes, multiple trainings

T02 - HOW LONG AGO WAS THE LAST TRAINING YOU RECEIVED / PARTICIPATED IN? *

- Within the last week
- Within the last month
- Within the last two months
- More than two months ago

T03 - WHO PROVIDED TRAINING(S)? *

*multiple responses accepted*

- Government
- Your place of practice / work
- NGOs
- Other, specify

OTHER TRAINING SOURCES, SPECIFY

T04 - ON WHICH OF THE FOLLOWING TOPICS HAVE YOU BEEN TRAINED IN RELATION TO COVID-19?

| | No | Yes |
|---|---|---|
| 01 - TRANSMISSION * | | |
| 02 - SYMPTOMS * | | |
| 03 - TESTS / DIAGNOSTIC * | | |
| 04 - ISOLATION / QUARANTINE * | | |
| 05 - PREVENTION * | | |
| 06 - CARE FOR PATIENTS * | | |
| 07 - COVID-19 TREATMENTS CENTERS * | | |
| 08 - OUTREACH * | | |

#### PROTECTIVE EQUIPMENT

PE01 - AT THIS TIME, HOW AVAILABLE ARE PROTECTIVE EQUIPMENT LIKE MASKS OR GLOVES AT YOUR PLACE OF PRACTICE / WORK? *

*in the last week*

Not at all available — Little availability — Somewhat available — Very available

PE02 - AND DURING NORMAL OPERATIONS, BEFORE COVID-19, HOW AVAILABLE ARE SUCH PROTECTIVE EQUIPMENT? *

Not at all available — Little availability — Somewhat available — Very available

PE03 - COMPARED TO NORMAL OPERATIONS, HOW FREQUENTLY DO YOU CURRENTLY USE PROTECTIVE EQUIPMENT? *

*in the last 4 weeks*

Much more frequently — More frequently — Same frequency — Less frequently — Much less frequently

PE04 - AND CURRENTLY, WHEN SEEING PATIENTS, HOW FREQUENTLY DO YOU WEAR MASKS? *

*in the last 4 weeks*

All the time — Most of the time — Some of the time — Rarely — Never — (Never see patients)

PE05 - WHAT FACTORS, IF ANY, EXPLAIN WHY YOU MAY NOT WEAR A MASK WHEN SEEING PATIENTS *

*multiple responses accepted*

- ☐ Masks not always available
- ☐ Masks of poor quality
- ☐ Masks are uncomfortable
- ☐ Masks are ineffective
- ☐ Patient presents no risk
- ☐ Other, specify

OTHER FACTORS, SPECIFY

PE06 - HOW DO YOU GET MASKS FOR YOUR WORK? *

*multiple responses accepted*

- ☐ From the place of care / work
- ☐ From NGOs
- ☐ Personal purchase
- ☐ Gifts from friends / family

#### TESTING AND CARE

TC01 - DO YOU CURRENTLY SEE OR INTERACT WITH PATIENTS POTENTIALLY INFECTED WITH THE VIRUS THAT CAUSES COVID-19? *

◯ No ◯ Yes

TC02 - HOW OFTEN DO THE PATIENTS YOU SEE OR INTERACT WITH WEAR MASKS? *

*in the last 4 weeks*

◯ All the time ◯ Most of the time ◯ Some of the time ◯ Rarely ◯ Never ◯ (Never see patients)

TC03 - HOW DO YOU JUDGE YOUR ABILITY TO DETECT POTENTIAL CASES OF COVID-19 AMONG PATIENTS YOU INTERACT WITH? *

◯ Not at all able ◯ Little able ◯ Somewhat able ◯ Very able

TC04 - AND IN YOUR OPINION, HAVE YOU SEEN OR INTERACTED WITH PATIENTS WHO SHOWED SYMPTOMS CONSISTENT WITH COVID-19? *

◯ No ◯ Yes

TC05 - HAVE YOU PERSONALLY SOUGHT TO OBTAIN TESTING FOR PATIENTS SHOWING SYMPTOMS CONSISTENT WITH COVID-19? *

*This is testing for patients*

◯ No ◯ Yes

TC06 - GENERALLY, WHAT BARRIERS EXIST TO ACCESSING TESTS FOR PATIENTS, IN YOUR OPINION? *

*multiple responses accepted*

- ☐ Testing supplies are unavailable
- ☐ Not prioritized for testing by current testing policies
- ☐ Did not exhibit COVID-19 symptoms
- ☐ Lack of transportation
- ☐ Restriction in mobility
- ☐ Financial cost of testing
- ☐ Insurance does not cover testing
- ☐ Extended wait times for testing
- ☐ Concerns of contracting COVID-19 (e.g. poor social distancing measures implemented at testing site)
- ☐ Other; Specify

OTHER BARRIERS, SPECIFY

#### PERSONAL RISK AND RESILIENCE

PRR01 - IN YOUR OPINION, WHAT IS YOUR OWN LEVEL OF RISK TO CONTRACT COVID-19? *

Not at all at risk | Little risk | Somewhat a risk | Very much at risk

PRR02 - TO WHAT EXTENT ARE YOU CONCERNED ABOUT TRANSMITTING COVID-19 TO PATIENTS / HOSPITAL VISITORS? *

Not at all concerned | Little concerned | Somewhat concerned | Very concerned

PRR03 - TO WHAT EXTENT ARE YOU CONCERNED ABOUT TRANSMITTING COVID-19 TO YOUR FRIENDS / FAMILY? *

Not at all concerned | Little concerned | Somewhat concerned | Very concerned

PRR04 - HOW DO YOU JUDGE YOUR CONTRIBUTION TOWARDS REDUCING THE SPREAD OF COVID IN YOUR COMMUNITY? *

Not at all able | Little able | Somewhat able | Very able

PRR05 - OVER THE LAST TWO WEEKS, HOW OFTEN HAVE YOU BEEN BOTHERED BY FEELING NERVOUS, ANXIOUS OR ON EDGE? *

Nearly every day | More than half of the days | Several days | Not at all

PRR06 - OVER THE LAST TWO WEEKS, HOW OFTEN HAVE YOU BEEN BOTHERED BY NOT BEING ABLE TO STOP OR CONTROL WORRYING? *

Nearly every day | More than half of the days | Several days | Not at all

PRR07 - HAVE YOU EXPERIENCED ANY ANGER/VERBAL OR OTHER ABUSE FROM COMMUNITY MEMBERS OR ACQUAINTANCES BECAUSE OF YOUR PROFESSION AS A RESULT OF COVID-19? *

- ○ No
- ○ Yes

PRR08 - PLEASE SPECIFY WHAT KIND OF ABUSE YOU HAVE EXPERIENCED FROM COMMUNITY MEMBERS OR ACQUAINTANCES BECAUSE OF YOUR PROFESSION SINCE THE PANDEMIC BEGAN. *

*multiple responses accepted*

- ☐ Insults
- ☐ Being forced out of my housing / rental
- ☐ Threats of physical violence
- ☐ Sexual harassment
- ☐ [illegible]

#### SURVEY INFORMATION

HOW DID YOU HEAR ABOUT THIS SURVEY?

- [ ] Employer
- [ ] Medical colleagues
- [ ] Government communication
- [ ] Professional Association
- [ ] NGOs / UN agencies
- [ ] Relatives and friends
- [ ] Email
- [ ] Twitter
- [ ] Facebook
- [ ] Other; specify

OTHER, SPECIFY
